# Investigating a possible causal relationship between maternal serum urate concentrations and offspring birthweight: A Mendelian randomization study

**DOI:** 10.1101/2022.02.28.22271245

**Authors:** Caitlin S. Decina, Rhian Hopkins, Jack Bowden, Beverly M. Shields, Deborah A. Lawlor, Nicole M. Warrington, David M. Evans, Rachel M. Freathy, Robin N. Beaumont

## Abstract

**Background:** Higher urate levels associate with higher systolic blood pressure (SBP) in adults, and in pregnancy, with lower offspring birthweight. Mendelian randomization (MR) analyses suggest a causal effect of higher urate on higher SBP and of higher maternal SBP on lower offspring birthweight. If urate causally reduces birthweight, it might confound the effect of SBP on birth weight. We therefore tested for a causal effect of maternal urate on offspring birthweight.

**Methods:** We tested the association between maternal urate levels and offspring birthweight using multivariable linear regression in UK Biobank (UKB; *n*=133,187) and Exeter Family Study of Childhood Health (EFSOCH; *n*=872). We conducted two-sample MR to test for a causal effect of maternal urate (114 single nucleotide polymorphisms [SNPs]; *n*=288,649 European-ancestry) on offspring birthweight (*n*=406,063 European-ancestry; maternal SNP effect estimates adjusted for fetal effects). Using one-sample MR (*n*=199,768 UKB women), we also tested for a causal relationship between urate and SBP.

**Results:** Higher maternal urate was associated with lower offspring birthweight in UKB (28g lower birthweight per 1-SD higher urate [95% CI: -31, -25]; *P*=1.8×10^−75^), with a similar effect estimate in EFSOCH (22g [95%CI: -50, 6]; *P*=0.13). The MR causal effect estimate was directionally consistent, but smaller (−11g [95% CI: -25, 3]; *P*_IVW_=0.13). In women, higher urate was causally associated with higher SBP (1.7 mmHg higher SBP per 1-SD higher urate [95% CI: 1.4, 2.1]; *P*=7.8×10^−22^) consistent with that previously published in women and men.

**Conclusions:** The marked attenuation of the MR result of maternal urate on offspring birthweight, compared to the multivariable regression result suggests previous observational associations may be confounded. The 95% CIs of the MR result included the null but suggest a possible weak effect on birthweight. Maternal urate levels are unlikely to be an important contributor to offspring birthweight.

**Key Messages:**

- Previous research suggests higher maternal serum urate in pregnancy is associated with lower offspring birthweight, and Mendelian randomization studies suggest a causal relationship between urate and systolic blood pressure (SBP), and SBP and birthweight; a causal effect of urate on birthweight has not yet been estimated, and thus it is also unknown whether it confounds maternal SBP-birthweight effects.
- The causal effect estimate of urate on offspring birthweight was directionally consistent, but weaker than, observational estimates; the estimate had 95% confidence intervals which included the null.
- This study confirmed a causal association between serum urate and higher SBP in women consistent with that published from a sample of both women and men.
- Maternal urate is unlikely to be a major determinant of birthweight or an important confounder of the causal relationship between SBP and lower birthweight.

## Introduction

Lower birthweights are associated with adverse maternal and fetal outcomes in the perinatal period and in later life, most notably with conditions such as type two diabetes and cardiovascular diseases^1,2^. Various maternal factors are known to be associated with birthweight in observational studies, but the causal nature of the associations is not always known. The study of the genetics of cardiometabolic risk factors has led to clearer understanding of some of these associations, such as for systolic blood pressure (SBP), where higher maternal SBP has been causally associated with lower offspring birthweight^3–5^.

Higher maternal serum urate levels are associated with lower offspring birthweight in epidemiological studies^6,7^ and are associated with higher SBP in pregnancies with low birthweight outcomes^8^. Raised serum urate levels are associated with cardiovascular diseases such as stroke, hypertension, atherosclerosis, and coronary heart disease in the general population^9–12^, and recently Mendelian randomization (MR) and clinical trial data of urate-lowering treatment have suggested that increases in serum urate exert a causal effect on higher SBP^13^. Although there is an apparent causal relationship between urate and SBP, and SBP and birthweight, a causal effect of urate on birthweight has not yet been estimated.

Observational associations in traditional epidemiological studies are prone to bias, confounding and reverse causality, and thus findings can be misleading. To overcome this, methods such as MR, which uses genetic variants as instrumental variables (IVs) to estimate the causal effect of the exposure of interest on the outcomes, can be applied to observational data^14^. Genetic variants can be regarded as valid IVs if three key assumptions are met: (i) the IVs are robustly associated with the exposure of interest, (ii) there are no residual confounders between the genetic IVs and outcome, and (iii) the IVs are (potentially) associated with the outcome only through the exposure of interest^15^. Indeed, MR studies share many similarities with randomized controlled trials. As alleles randomly segregate and assort independently, variants that proxy exposures should also be independent of environmental influences that might otherwise induce confounding between the exposure and outcome^14^. Also, because an individual’s germline genotype must precede the outcome of interest, reverse causality is typically not an issue in MR studies. MR studies have recently been applied to examine potential causal relationships between maternal environmental exposures and their offsprings’ outcomes^3–5^.Methods have been developed within this context to enable two-sample MR, i.e. where different samples of individuals are used to estimate the maternal variant-maternal exposure relationship and the maternal variant-offspring outcome relationship^16,17^. These methods are useful for incorporating data from large consortia and thus increasing statistical power^18,19^.

In this study, we aimed to (i) explore associations between maternal serum urate and offspring birthweight in a cohort of pregnant women from the Exeter Family Study of Childhood Health (EFSOCH), as well as with retrospective maternally-reported offspring birthweight in women from the UK Biobank (UKB), (ii) use MR to test for a causal effect of maternal urate concentration on offspring birthweight, and (iii) use MR to confirm that urate is causally related to SBP in women.

## Methods

### Study Overview

We first estimated the observational association between maternal serum urate and offspring birthweight in a study of 872 pregnant women and their babies in EFSOCH^20^, and in 133,187 women from UKB^21^, and compared these with previously published associations. We compared our urate-birthweight observational associations with the causal effect estimate obtained from univariate two-sample MR analyses using a set of associated genetic variants, or single nucleotide polymorphisms (SNPs), as instrumental variables for maternal serum urate. We used estimates for the variant association with the exposure and outcome from the largest, most recent genome-wide association studies (GWAS) available for each variable^4,22^. We then tested for a causal relationship between serum urate and SBP in women only by performing one-sample MR in 199,768 women within the UKB to compare to recently published findings obtained from a sample of both men and women. All datasets used comprised participants of genetically defined European ancestry to obtain a large, well-powered sample while ensuring consistent allele frequencies across datasets. Figure 1 depicts the putative relationships between all variables considered, with the relationship between maternal urate and offspring birthweight being the primary focus of the investigation.

**Figure 1.**
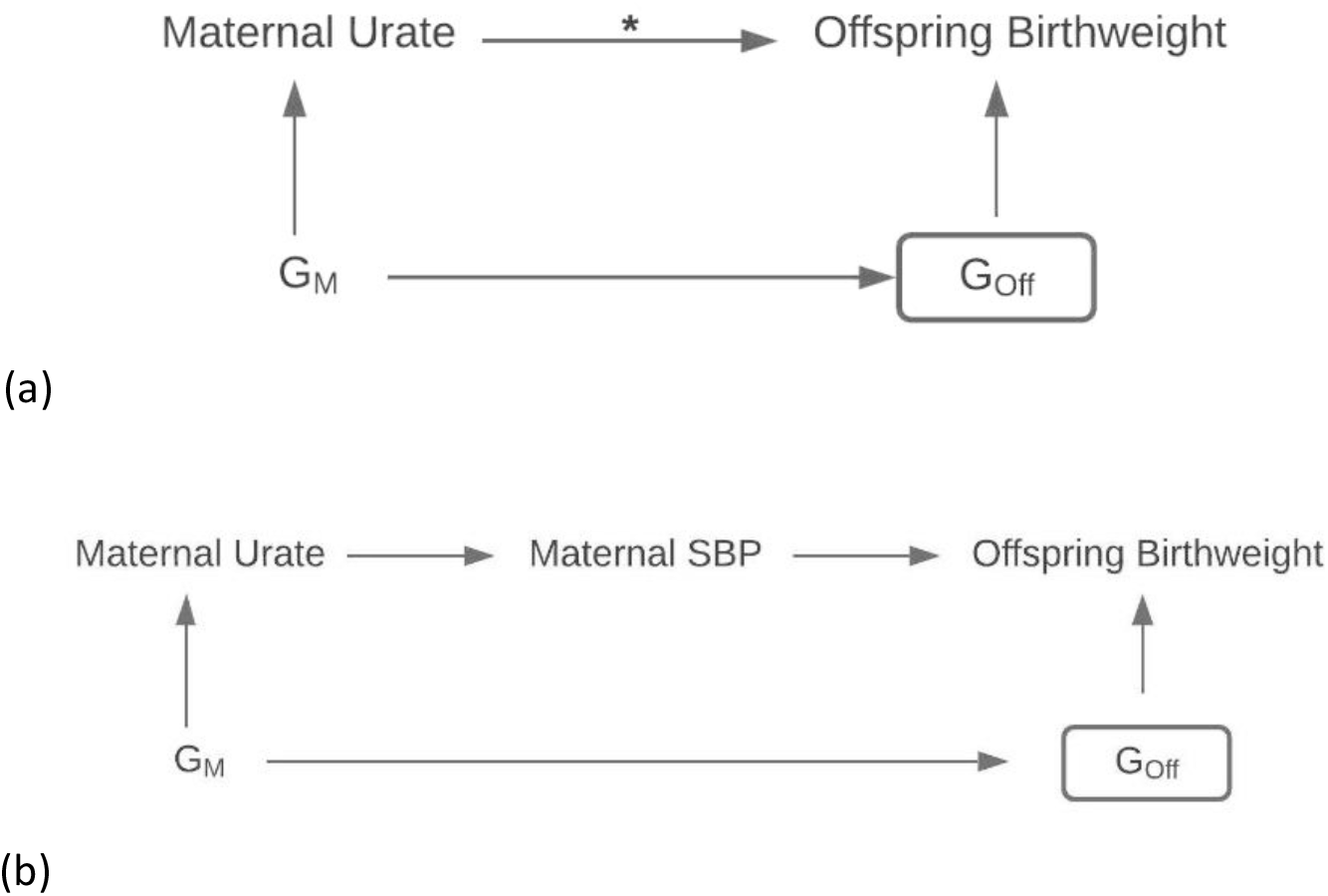
Directed acyclic graph illustrating the plausible (a), or known (b), relationships among key variables (solid arrows; e.g. Mendelian randomization (MR) analyses show a causal effect of higher urate resulting in higher systolic blood pressure (SBP) and higher SBP resulting in lower birthweight), where offspring birthweight is the outcome, and maternal urate is the exposure of interest. (a) The putative relationships of the main model we aimed to investigate, estimating the total causal effect of maternal urate on offspring birthweight where the asterisk denotes the causal association being tested. Multivariable observational analyses suggest an inverse effect of higher maternal circulating urate lowering infant birthweight. We used MR with maternal genetic instrumental variables (G_M_) to explore whether there was an inverse total causal effect of urate on birthweight, and if so, whether urate might be a confounder of the SBP-birthweight inverse effect. A rectangular box denotes the variable and its effects have been adjusted for. As offspring genome (G_Off_) has effects on own birthweight, maternal effects were estimated as independent of the offspring genome to capture the maternal genome’s effect on offspring birthweight alone. (b) The known relationships between urate and SBP, and SBP and offspring birthweight. Our MR results confirm a similar magnitude of effect of urate on SBP in women and an inverse effect of urate on birthweight, with a magnitude (based on the point estimate) that would be consistent with this effect being totally mediated via SBP, hence urate is unlikely to be a confounder of the SBP-birthweight effect. SBP = systolic blood pressure; G_M_ = maternal genotype; G_Off_ = offspring’s genotype.

### Observational Association of Maternal Urate with Offspring Birthweight

We performed multivariable linear regression to explore the association between maternal urate levels measured at 28 weeks gestation (exposure of interest) and offspring birthweight (outcome) in a sample of 872 unrelated women within the EFSOCH study. We included maternal age, height, body mass index (BMI), smoking status, 28-week fasting glucose, estimated glomerular filtration rate (eGFR), and Townsend deprivation index (TDI), as well as child’s sex and gestational age as covariables in the association model. EFSOCH did not have data available for participants’ alcohol intake habits which therefore could not be adjusted for in this model.

The EFSOCH sample provided the benefit of investigating associations using measurements taken during pregnancy, although it was limited in sample size. We therefore also performed observational analyses in a larger sample of UKB women, with the limitation that participants’ urate levels were measured many years after pregnancy. A multivariable linear regression model was used to explore the association between maternal urate levels measured on average 31.6 (SD=10.1) years after delivery and offspring birthweight among 133,187 unrelated women within the UK Biobank who reported the birthweight of their first child. This model was also adjusted for maternal age at study enrolment (when urate was measured), maternal age at first birth, height, BMI, smoking status, alcohol intake, eGFR, blood glucose level, TDI, and assessment centre location. Information on child’s sex and gestational age at delivery were not available in UKB and therefore were not adjusted for in this analysis.

### Genetic Instruments for Maternal Serum Urate Concentration

The genetic instruments used for maternal serum urate in the MR analysis were genetic variants identified in a recently published trans-ancestry GWAS of serum urate in 457,690 individuals where UKB was not used for discovery analysis of urate-associated variants^22^. This GWAS identified a total of 183 loci that contained at least one genome-wide significant SNP (*P*≤5×10^−8^), for which the index SNPs at these 183 loci explained 7.7% of variance in serum urate levels. For our study, we restricted this list to include only variants for which *P*<5×10^−8^ in the European ancestry-specific meta-analysis (*n*=288,649), resulting in 114 independent SNPs (Supplementary Table S1).

### Validating Genetic Instrument for Urate in Pregnant Women

As the urate-associated SNPs used in this study were identified in a sample of males and non-pregnant females, we wanted to determine whether the genetic instruments were good predictors of urate levels in pregnancy as well as outside of pregnancy. Therefore, we created a weighted, standardized genetic score (GS) for urate and tested its association with urate levels in women in the EFSOCH study where urate was measured in participants in both periods: at 28 weeks gestation (*n*=872) and approximately six years post-pregnancy (*n*=470)^20^. The GS was calculated according to Equation (1),

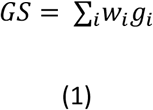

where w_*i*_ is the weight for SNP *i* and g_*i*_ is the genotype (number of effect alleles 0-2) at SNP *i*. The SNP weightings were the regression coefficients obtained from the most recently reported GWAS of urate as mentioned above^22^. The calculated GS was then standardized and the urate levels for participants (µmol/L) in each time period regressed on the standardised genetic score.

Of the 114 urate-associated SNPs, 10 did not have genotype data available in EFSOCH. For these 10 SNPs, we searched for suitable proxies using the LDproxy tool^23^. SNPs found with the highest R^2^ (minimum R^2^=0.8) in a European population, within a 500kbp window of the target SNP, were selected for analysis. Four SNPs did not have proxies available which met the R^2^ threshold and were excluded from analysis, therefore, a total of 110 urate-associated SNPs were used to compose the GS (Supplementary Material Table S2).

### Mendelian Randomization of Maternal Urate on Offspring Birthweight

We used two-sample MR to test for a causal association between maternal urate and offspring birthweight. The variant-exposure associations were obtained using the European-ancestry-specific summary data from the serum urate GWAS mentioned previously (Sample 1)^22^. The variant-outcome associations were obtained from a GWAS of own birthweight and offspring birthweight in a total of 406,063 European-ancestry individuals from the Early Growth Genetics (EGG) Consortium and UKB, where maternal genetic effects on birthweight were estimated approximately conditional on fetal effects using a weighted linear model (WLM) (Sample 2)^4^. The total number of participants were comprised of 101,541 UKB participants who reported their own birthweight and the birthweight of their first child, 195,815 UKB and EGG participants with own birthweight data, and 108,707 UKB and EGG participants with offspring birthweight data.

Inverse variance weighted (IVW) MR was performed along with additional pleiotropy-robust sensitivity analyses, including MR-Egger, weighted median, and penalised weighted median estimators. The IVW method of estimating causal effects is commonly used for individual and summary level data^24^, while MR-Egger provides a means to test and correct for bias in the IVW estimate when variants affect the outcome through pathways other than the exposure of interest (pleiotropy)^25^. Weighted median MR additionally provides a reliable estimate when up to half of the variance explained in the genetically predicted exposure comes from invalid IVs^26^, with penalization providing added robustness to large outliers. Details of the R code for the MR analyses are provided elsewhere^25,26^.

### Validating a Causal Effect of Urate on Systolic Blood Pressure

We sought to confirm a causal relationship between serum urate and SBP in UKB women, our main population of interest, as previous causal findings used a mixed sex sample excluding UKB participants^13^. We thus performed one-sample MR in 199,768 women within the UKB using two-stage least squares, with estimation conducted using the **ivreg2** function in Stata version 16.0^27,28^.

## Results

### Observational Associations

Baseline characteristics of the EFSOCH and UKB women included in each multivariable linear regression model are described in Tables 1 and 2, respectively.

**Table 1.**
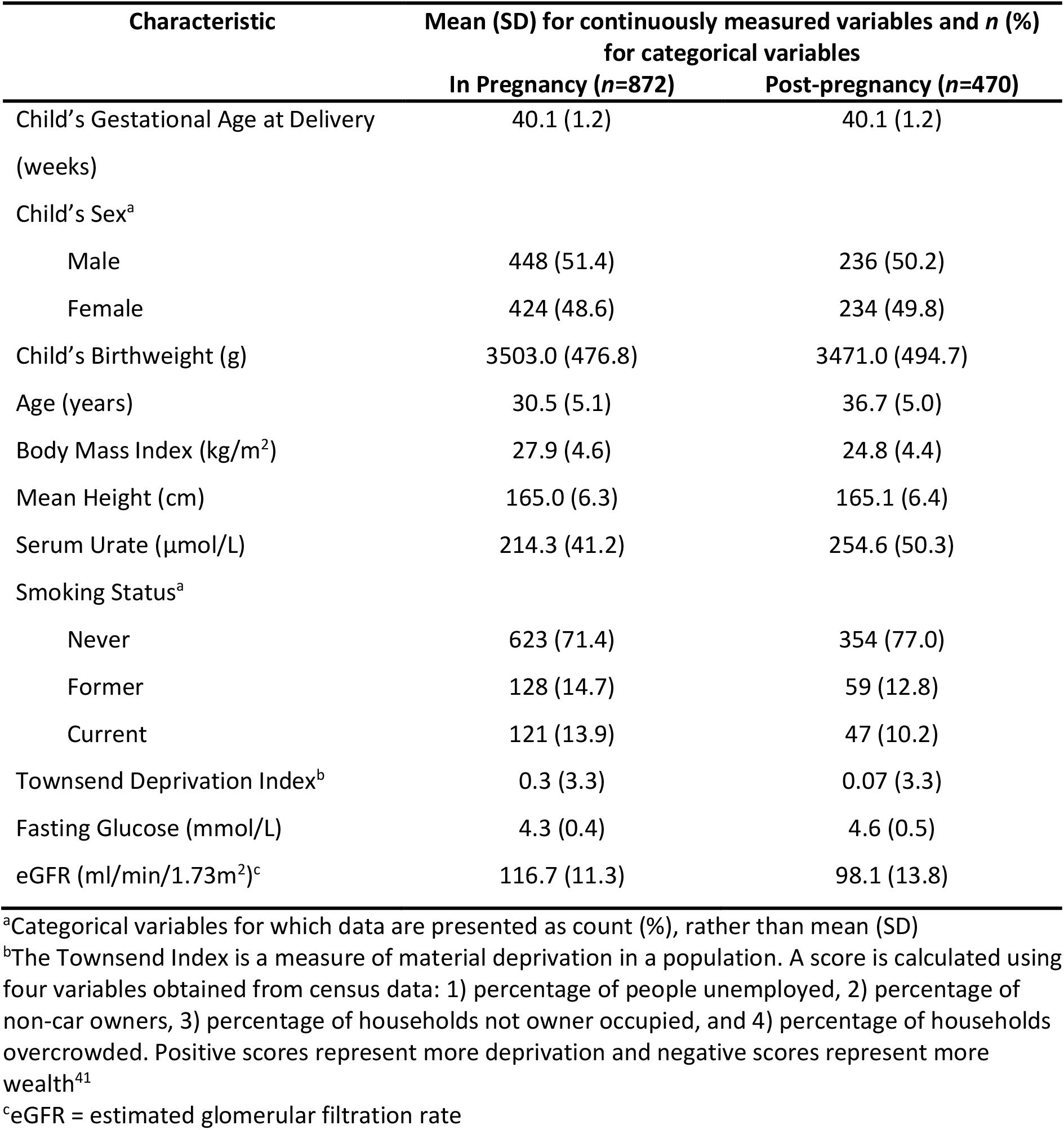
Descriptive characteristics of Exeter Family Study of Childhood Health women included in the multivariable linear model of factors associated with child’s birthweight, both in pregnancy and postpartum. Measurements during pregnancy were taken at 28 weeks gestation.

| Characteristic | Mean (SD) for continuously measured variables and *n* (%)<br>for categorical variables |  |
| --- | --- | --- |
|  | In Pregnancy (*n*=872) | Post-pregnancy (*n*=470) |
| Child's Gestational Age at Delivery<br>(weeks) | 40.1 (1.2) | 40.1 (1.2) |
| Child's Sex<sup>a</sup> |  |  |
| Male | 448 (51.4) | 236 (50.2) |
| Female | 424 (48.6) | 234 (49.8) |
| Child's Birthweight (g) | 3503.0 (476.8) | 3471.0 (494.7) |
| Age (years) | 30.5 (5.1) | 36.7 (5.0) |
| Body Mass Index (kg/m<sup>2</sup>) | 27.9 (4.6) | 24.8 (4.4) |
| Mean Height (cm) | 165.0 (6.3) | 165.1 (6.4) |
| Serum Urate (μmol/L) | 214.3 (41.2) | 254.6 (50.3) |
| Smoking Status<sup>a</sup> |  |  |
| Never | 623 (71.4) | 354 (77.0) |
| Former | 128 (14.7) | 59 (12.8) |
| Current | 121 (13.9) | 47 (10.2) |
| Townsend Deprivation Index<sup>b</sup> | 0.3 (3.3) | 0.07 (3.3) |
| Fasting Glucose (mmol/L) | 4.3 (0.4) | 4.6 (0.5) |
| eGFR (ml/min/1.73m<sup>2</sup>)<sup>c</sup> | 116.7 (11.3) | 98.1 (13.8) |

<sup>a</sup>Categorical variables for which data are presented as count (%), rather than mean (SD)
<sup>b</sup>The Townsend Index is a measure of material deprivation in a population. A score is calculated using four variables obtained from census data: 1) percentage of people unemployed, 2) percentage of non-car owners, 3) percentage of households not owner occupied, and 4) percentage of households overcrowded. Positive scores represent more deprivation and negative scores represent more wealth<sup>41</sup>
<sup>c</sup>eGFR = estimated glomerular filtration rate

**Table 2.** Baseline characteristics of UK Biobank women included in the multivariable model of factors associated with first child’s birthweight (*n*=133,187).

| Characteristic | Mean (SD) for continuously measured variables<br>and $n$ (%) for categorical variables |
| --- | --- |
| Child's birthweight (g) | 3227.8 (477.1) |
| Age at enrolment (years) | 57.7 (7.7) |
| Age at first birth (years) | 26.0 (5.0) |
| Body Mass Index ($\text{kg}/\text{m}^2$) | 27.0 (5.0) |
| Height (cm) | 162.5 (6.1) |
| Serum Urate ($\mu\text{mol}/\text{L}$) | 270.8 (65.4) |
| Smoking Status<sup>a</sup> |  |
| Never | 78,214 (58.7) |
| Former | 44,337 (33.3) |
| Current | 10,636 (8.0) |
| Alcohol Intake Frequency<sup>a</sup> |  |
| Daily or almost daily | 22,143 (16.6) |
| 3 or 4 times/week | 28,766 (21.6) |
| 1 or 2 times/week | 35,594 (26.7) |
| 1-3 times/month | 17,321 (13.0) |
| Special occasions only | 18,928 (14.2) |
| Never | 10,435 (7.8) |
| Townsend Deprivation Index | -1.7 (2.8) |
| Blood Glucose ($\text{mmol}/\text{L}$) | 5.0 (0.7) |
| eGFR ($\text{ml}/\text{min}/1.73\text{m}^2$)<sup>b</sup> | 89.9 (12.9) |

<sup>a</sup>Categorical variables for which data are presented as count (%), rather than mean (SD)
<sup>b</sup>eGFR = estimates glomerular filtration rate

Multivariable modelling using the sample of EFSOCH women indicated there was weak evidence of an association between maternal urate levels and offspring birthweight, with a wide 95% confidence interval due to a relatively small sample size (22g lower birthweight per 1-SD higher maternal urate [95% CI: -50, 6]; *P*=0.13; Figure 2 and Supplementary Table S3).

**Figure 2.**
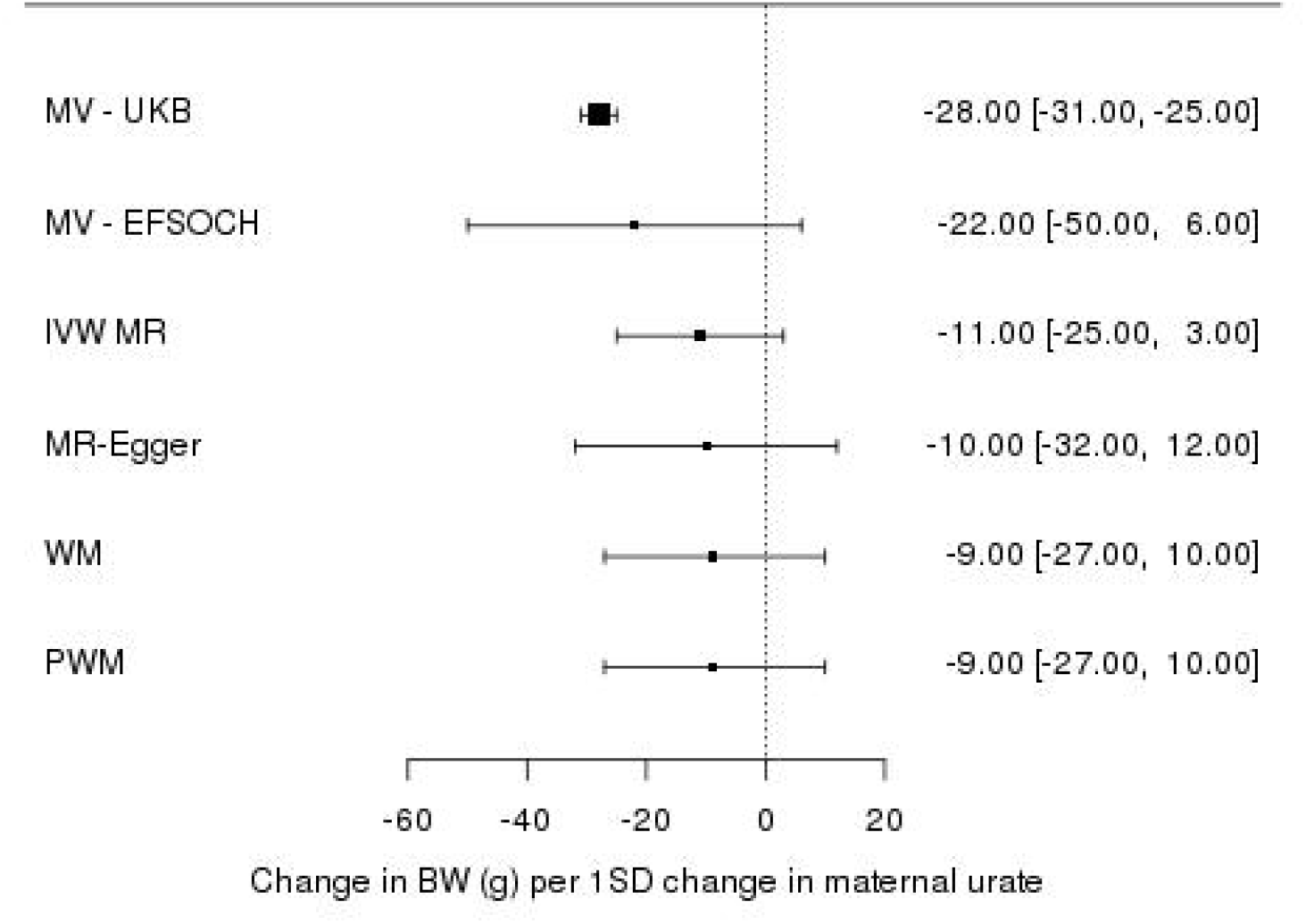
Forest plot of the effect of maternal serum urate on offspring birthweight: observationally using participants in the UK Biobank (*n*=133,187), observationally using participants in the Exeter Family Study of Childhood Health (*n*=872), and using univariate Mendelian randomization (inverse variance weighted method) with additional sensitivity methods. Each estimate is the change in birthweight in grams per 1 SD change in urate level [95% CI]. MV=multivariable linear regression; UKB=UK Biobank; EFSOCH=Exeter Family Study of Childhood Health; IVW=inverse variance weighted; MR=Mendelian randomization; WM=weighted median; PWM=penalised weighted median; BW=birthweight; SD=standard deviation.

The same associations were then investigated in a much larger sample of UKB women, with the caveat of urate having been measured at a mean of 31.6 (SD=10.1) years post-pregnancy. Here, results indicated that higher maternal serum urate concentration was associated with a lower offspring birthweight (28g lower birthweight of first child per 1-SD higher maternal urate [95% CI: -31, -25]; *P*=1.8×10^−75^), consistent with that found in EFSOCH (Figure 2 and Supplementary Table S4).

### Association between Urate Genetic Score and Urate Measured During and Post Pregnancy

In the sample of EFSOCH women, the GS for urate showed similar association with urate levels measured in pregnancy at an average maternal age of 30.3 (SD=5.3) years compared to post-pregnancy measured at an average maternal age of 36.7 (SD=5.0) years (13.1 µmol/L [95% CI: 10.4, 15.8] and 13.4 µmol/L [95% CI: 9.0, 17.8] higher urate per 1-SD higher urate GS, respectively), indicating that the SNPs associated with urate levels in the general population used for the MR analyses are similarly associated with urate levels in a population of pregnant women (Figure 3).

**Figure 3.**
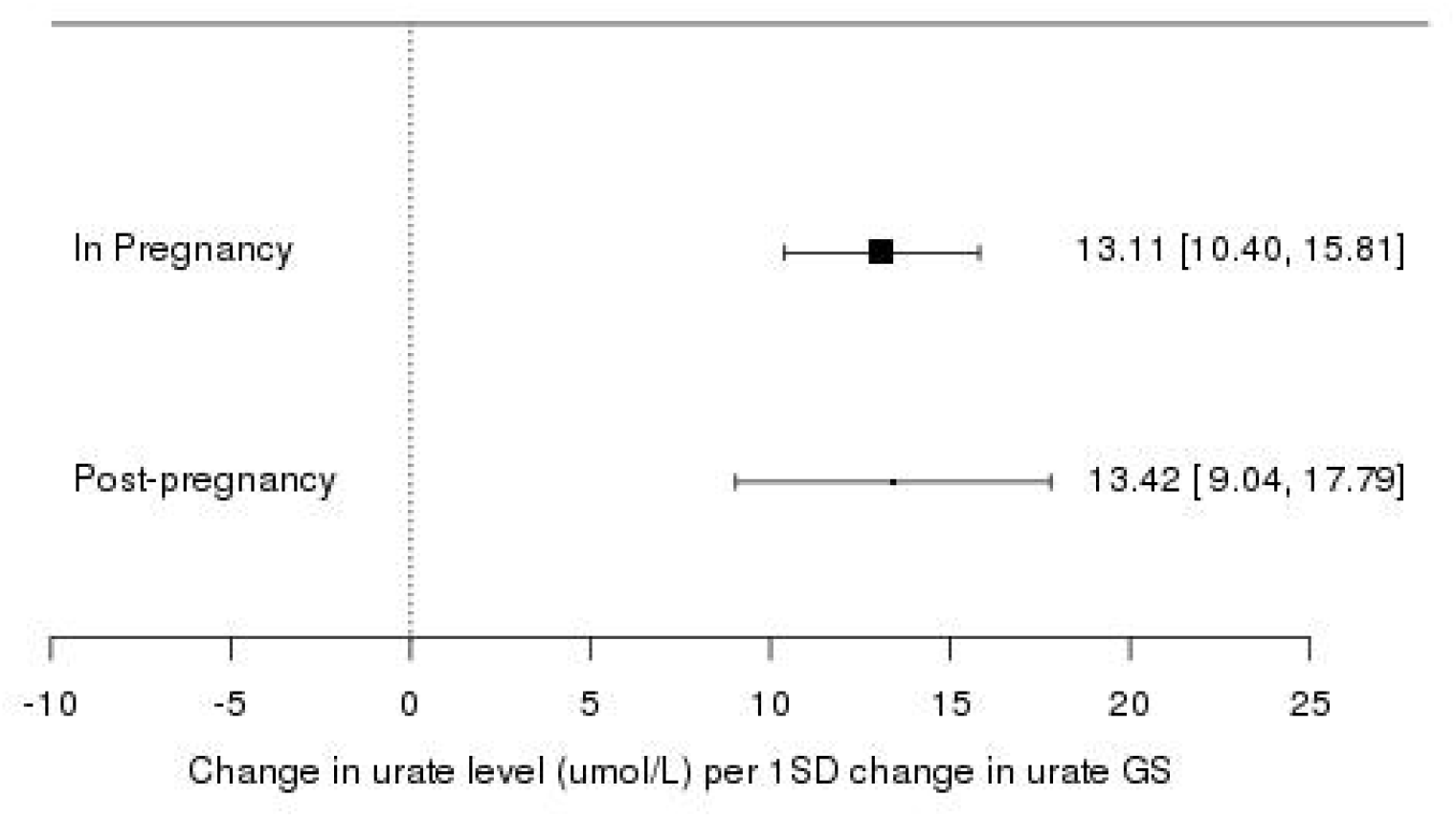
Forest plot of the association between a 110-SNP urate genetic score and urate levels measured in pregnancy (*n*=872) and post-pregnancy (*n*=470) in the Exeter Family Study of Childhood Health. Each estimate is the change in urate level (µmol/L) per 1 SD change in urate genetic score [95% CI]. SD=standard deviation; GS=genetic score.

### Two-sample Mendelian Randomization

We did not find strong evidence for a causal effect of maternal urate level on offspring birthweight, though the causal effect estimate for urate was directionally consistent with the observational estimate (11g lower birthweight per 1-SD higher urate [95% CI: -25, 3]; *P*_IVW_=0.13). Sensitivity analyses showed similar results (Figure 2 and Supplementary Figure S1).

### Causal Effect of Urate on Systolic Blood Pressure

Within UKB women, we found strong evidence for a causal effect of serum urate levels on SBP (1.7 mmHg higher SBP per 1-SD higher serum urate [95% CI: 1.4, 2.1]; *P*=7.8×10^−22^]). Based on 1-SD of SBP = 24.23 mmHg, this is equivalent to 0.07 SD higher SBP per 1-SD higher urate.

## Discussion

In our own investigation we found 22g and 28g lower birthweight per 1-SD higher maternal urate in observed confounder adjusted multivariable regression analyses in the EFSOCH and UKB studies, respectively. This is consistent with previous observational studies which have found that higher maternal urate levels show an association with lower offspring birthweight^6,7^. When investigating a causal association of maternal urate on offspring birthweight using MR, the effect estimate was directionally consistent with the observational estimates, though imprecise and including the null. The association between maternal SBP and lower birthweight, has previously been established as causal, e.g. by Tyrrell et al. and Warrington et al.^3,4^. Recently published MR and clinical trial data has now provided evidence of a causal effect of serum urate on SBP^13^. When investigating this causal relationship in UKB women only, our result was consistent with their findings (0.07 vs. 0.09 SD units of SBP per 1-SD higher serum urate, respectively).

Given the estimated causal effect of urate on SBP found here in women, we would correspondingly expect approximately 12g lower birthweight per 1-SD higher maternal urate concentration if all of the effect of urate was mediated by SBP (Supplementary Note). This estimated difference in birthweight sits within the 95% CIs of the urate on birthweight MR causal effect estimate and is indeed very close to the effect estimate itself of approximately 11g lower birthweight per 1-SD change in urate. Thus, our findings suggest that urate has at most a modest effect on birthweight that is fully mediated by SPB, suggesting that it is unlikely to be a key confounder of the maternal SBP-offspring birthweight effect (Figure 1b). The span of 95% CIs of the MR analyses cannot rule out a small direct causal effect, and potentially some confounding of the SBP birthweight effect.

Serum urate, or uric acid, is thought to be involved in the biological processes of oxidative stress and inflammation, with particular action in endothelial cells causing vascular damage, mechanisms which may explain its role in increasing SBP and the development of cardiovascular diseases ^29,30^. With little evidence to currently support a direct causal role for maternal urate lowering mean offspring birthweight, it is possible that higher mean urate levels seen in pregnancies with small babies reflect the impact of higher urate on blood pressure and of higher blood pressure on lower birthweight. Maternal pre-existing high blood pressure, and hypertensive disorders of pregnancy, are related to placental vascular malperfusion and dysfunction, which may then exert greater impact on fetal growth than urate might influence placental function through direct mechanisms^31^. A pathogenic role for urate has also been suggested in the development of pre-eclampsia^32–34^, a hypertensive disorder of pregnancy characterised by the onset of high blood pressure and substantial proteinuria ≥ 20 weeks’ gestation, for which high urate levels and low offspring birthweight outcomes are commonly associated^35–37^. The finding in the current study of no strong evidence for a causal effect of maternal urate level on offspring birthweight is therefore not entirely unexpected given that other circulatory system-related factors, such as SBP and abnormal vascular changes in the placenta seen in pre-eclampsia cases, have already been shown to play a role in low birthweight outcomes^3,31,38^. Investigating the role of maternal urate in pre-eclampsia using causal methods indeed presents an avenue for future research and an opportunity to provide further clarity on how urate may influence birthweight along a causal pathway.

There are some limitations worth noting in this study. As mentioned previously, the use of the UKB as a population sample is limited due to participants being older at baseline and not having measurements specific to pregnancy. The UKB does, however, provide the significant advantage of being large and the most well-powered sample available for this investigation. To ensure that our findings would not be unduly influenced by these demographic differences, we used EFSOCH to test both the association of urate on offspring birthweight observationally, and the association between the urate SNPs used and urate levels in women who were assessed in pregnancy. These comparisons yielded consistent results across cohorts and time periods, therefore we do not expect use of the UKB in our analyses to be problematic.

Additionally, there is a small overlap between the cohorts that contributed to the GWAS of serum urate and the EGG consortium GWAS of own (fetal) birthweight, but not with the maternal GWAS of offspring birthweight. With respect to impact this might have for overlap between SNP-exposure estimates and SNP-outcome estimates in our study, there will be some influence on the WLM estimates via the fetal effect estimates that are included, but not via the maternal estimates, therefore only ∼50% overlap. Together, these cohorts make up a very small proportion of the fetal GWAS sample (*n*=5766/321,223; 1.7%), thus any risk of bias from inclusion of these studies will be minimal.

Lastly, the SNP-birthweight effect estimates used in our analyses were mostly unadjusted for gestational duration^4^. Evidence in the literature is mixed on whether maternal SBP affects fetal growth directly, or if the birthweight effect is mediated by an effect of maternal SBP on gestational duration^39^. Previous causal MR analyses suggest higher SBP has a direct effect on lower birthweight^3,4^, however, recent work suggests that increased maternal SBP is causally associated with shorter gestational duration^40^. As the birthweight associations used in this study are thus not fully adjusted for gestational age at delivery, we are aware that the effects on birthweight being tested here could reflect a combination of factors, including fetal growth rates, or gestational duration, or both.

As mentioned previously, we used the currently most well-powered sample available to investigate a causal relationship between maternal urate and birthweight, however, our conclusions may still be limited by sample size. While our results suggest that a large causal effect of urate on birthweight is unlikely, they do not rule out smaller effects. Further investigation will allow for greater precision in effect estimates and thus to determine whether small, but potentially biologically meaningful, effects exist especially in cases such as pre-eclampsia where the impact and role of urate is not confirmed, and where small changes could influence whether a pregnancy is classified as higher risk.

In conclusion, there was a lack of strong evidence for a causal effect of maternal urate on offspring birthweight, though 95% CIs suggest the possibility of weak influence that larger studies may provide a more precise estimate of in the future. Stronger associations seen in previously published observational studies may be confounded by other maternal physiological factors, such as the effect of SBP. Current evidence indicates that maternal SBP mediates any effect of urate on birthweight and that urate is unlikely to confound the causal effect of maternal SBP on offspring birthweight. Further research into the relationship between maternal urate and pre-eclampsia using causal methods could help further clarify a role for urate in pregnancy.

## Data Availability

The genotype and phenotype data are available on application from the UK Biobank (http://www.ukbiobank.ac.uk/). Individual cohorts participating in the EGG Consortium should be contacted directly as each cohort has different data access policies. GWAS summary statistics of birthweight are freely available via the EGG website (https://egg-consortium.org/).
Genome-wide summary statistics for urate used in this study are publicly available at the CKDGen Consortium via http://ckdgen.imbi.uni-freiburg.de.
Summary statistics from EFSOCH are available on request. Researchers interested in accessing the data are expected to send a reasonable request by sending an email to the Exeter Clinical Research Facility at.

https://egg-consortium.org/

http://ckdgen.imbi.uni-freiburg.de

## Ethics approval

This study was conducted using the UK Biobank resource, with access granted under project application number 7036. Details of patient and public involvement in the UK Biobank are available online (www.ukbiobank.ac.uk/about-biobank-uk/ and https://www.ukbiobank.ac.uk/wp-content/uploads/2011/07/Summary-EGF-consultation.pdf?phpMyAdmin=trmKQlYdjjnQIgJ%2CfAzikMhEnx6). No patients were specifically involved in setting the research question or the outcome measures, nor were they involved in developing plans for recruitment, design, or implementation of this study. No patients were asked to advise on interpretation or writing up of results. There are no specific plans to disseminate the results of the research to study participants, but the UK Biobank disseminates key findings from projects on its website.

Ethical approval for the Exeter Family of Childhood Health was given by the North and East Devon (UK) Local Research Ethics Committee (approval number 1104), and informed consent was obtained from the parents of the newborns.

The EGG consortium birthweight GWAS summary data used in this study did not require prior approval for access and is freely available online.

## Author contributions

C.S.D., J.B., N.M.W, D.M.E., R.N.B. and R.M.F contributed to study design. C.S.D. performed the analyses in this study, with contribution by R.H., and drafted the manuscript. Data interpretation and statistical analysis was aided by J.B., D.A.L., R.N.B, and R.M.F. Data curation of the EFSOCH study was performed by B.M.S. This work was jointly directed by R.N.B and R.M.F. All authors reviewed and edited previous versions of the manuscript. All authors read and approved the final manuscript. R.N.B. is the guarantor for the paper.

## Data availability

The genotype and phenotype data are available on application from the UK Biobank (http://www.ukbiobank.ac.uk/). Individual cohorts participating in the EGG Consortium should be contacted directly as each cohort has different data access policies. GWAS summary statistics of birthweight used in this study are freely available via the EGG website (https://egg-consortium.org/). Genome-wide summary statistics for urate used in this study are publicly available at the CKDGen Consortium via http://ckdgen.imbi.uni-freiburg.de.

Summary statistics from EFSOCH are available on request. Researchers interested in accessing the data are expected to send a reasonable request by sending an email to the Exeter Clinical Research Facility at.

## Supplementary data

Supplementary data are available online.

## Acknowledgements

This research has been conducted using the UK Biobank Resource. This work was carried out under UK Biobank project number 7036.

This study represents independent research supported by the National Institute of Health Research (NIHR) Exeter Clinical Research facility. The views expressed are those of the author(s) and not necessarily those of the NHS, the NIHR or the Department of Health and Social care.

The Exeter Family Study of Childhood Health (EFSOCH) was supported by South West NHS Research and Development, Exeter NHS Research and Development, the Darlington Trust and the Peninsula NIHR Clinical Research Facility at the University of Exeter. The opinions given in this paper do not necessarily represent those of NIHR, the NHS or the Department of Health. Genotyping of the EFSOCH study samples was funded by the Wellcome Trust and Royal Society [grant 104150/Z/14/Z]. We would like to acknowledge Andrew Hattersley as the principal investigator, and Bea Knight for her contribution to data collection, of the EFSOCH study.

The authors would like to acknowledge the use of the University of Exeter High-Performance Computing (HPC) facility in carrying out this work. We acknowledge use of high-performance computing (and/or long-read sequencing) funded by a MRC Clinical Research Infrastructure award [MRC Grant: MR/M008924/1].

## Abbreviations

SBP: systolic blood pressure
MR: Mendelian randomization
GWAS: Genome-wide association study
SNP: single nucleotide polymorphism
UKB: UK Biobank
EFSOCH: Exeter Family Study of Childhood Health
BMI: body mass index
TDI: Townsend Deprivation Index
eGFR: estimated glomerular filtration rate
LD: linkage disequilibrium
EGG: Early Growth Genetics Consortium
WLM: weighted linear model
IVW: inverse variance weighted

