## Supplementary Table 3 and 4, Supplementary Figure 1 for "Investigating a possible causal relationship between maternal serum urate concentrations and offspring birthweight: A Mendelian randomization study"

**Supplementary Tables and Figures**

**Supplementary Table S3. Multivariable linear model of associations with child’s birthweight using data from 872 unrelated women within the Exeter Family Study of Childhood Health (adj. R-squared: 0.3057)**. Maternal serum urate was measured at mean 30.4 years of age and at 28 weeks of gestation.

| **Explanatory Variable** | **Change in birthweight (g) per 1 SD change in maternal trait** | | **SE^a^** | ***P*-value** |
| --- | --- | --- | --- | --- |
| Gestational Age | 188.7 | 13.7 | | 3.3x10^-39^ |
| Child’s Sex^b^ |  |  | |  |
| Male | Referent |  | |  |
| Female | -106.9 | 27.0 | | 8.2x10^-5^ |
| Mother’s Age (years) | 4.8 | 16.1 | | 0.76 |
| Mother’s Body Mass Index (kg/m^2^) | 87.4 | 14.8 | | 4.9x10^-9^ |
| Mother’s Mean Height (cm) | 103.7 | 13.8 | | 1.7x10^-13^ |
| **Serum Urate (µmol/L)** | -21.8 | 14.4 | | 0.13 |
| Smoking Status^b^ |  |  | |  |
| Never | Referent |  | |  |
| Former | -31.6 | 39.6 | | 0.43 |
| Current | -273.1 | 42.0 | | 1.4x10^-10^ |
| Townsend Deprivation Index | 7.4 | 14.0 | | 0.60 |
| Fasting Glucose (mmol/L) | 77.8 | 14.8 | | 1.8x10^-7^ |
| eGFR (ml/min/1.73m^2^)^c^ | 34.1 | 15.7 | | 0.03 |

^a^SE = standard error

^b^Categorical variables for which the data is presented as change in birthweight in grams compared to the referent category

^c^eGFR = estimated glomerular filtration rate

**Supplementary Table S4. Multivariable linear model of associations with birthweight of first child using data from 133,187 unrelated women within the UK Biobank (adj. R-squared: 0.0450).** Maternal serum urate was measured at mean 31.6 years post-pregnancy.

| **Explanatory Variable** | **Change in birthweight (g) per 1 SD change in maternal trait** | **SE^a^** | ***P*-value** |
| --- | --- | --- | --- |
| Age at enrolment (years) | 11.8 | 1.5 | 1.1x10^-14^ |
| Age at first birth (years) | 8.9 | 1.4 | 9.0x10^-11^ |
| Height (cm) | 89.1 | 1.3 | *P* < 1x10^-254^ |
| Body Mass Index (kg/m^2^) | 50.7 | 1.5 | 1.0x10^-254^ |
| **Serum Urate (µmol/L)** | -28.3 | 1.5 | 1.8x10^-75^ |
| Smoking Status^b^ |  |  |  |
| Never | Referent |  |  |
| Former | -18.1 | 2.8 | 1.5x10^-10^ |
| Current | -60.4 | 5.0 | 3.9x10^-34^ |
| Alcohol Intake Frequency^b^ |  |  |  |
| Daily or almost daily | Referent |  |  |
| 3 or 4 times/week | 0.2 | 4.2 | 0.97 |
| 1 or 2 times/week | -13.0 | 4.1 | 1.5x10^-3^ |
| 1-3 times/month | -10.8 | 4.8 | 0.03 |
| Special occasions only | -32.6 | 4.8 | 7.1x10^-12^ |
| Never | -31.6 | 5.7 | 2.4x10^-8^ |
| Townsend Deprivation Index | -13.8 | 1.4 | 2.0x10^-22^ |
| Blood Glucose (mmol/L) | 5.9 | 1.3 | 6.8x10^-6^ |
| eGFR (ml/min/1.73m^2^)^c^ | 15.3 | 1.5 | 1.3x10^-23^ |
| Assessment Centre^b^ |  |  |  |
| Manchester | Referent |  |  |
| Oxford | -41.4 | 10.8 | 1.2x10^-4^ |
| Cardiff | -23.0 | 10.3 | 0.02 |
| Glasgow | 17.9 | 10.3 | 0.08 |
| Edinburgh | 4.8 | 10.4 | 0.64 |
| Stoke | -46.1 | 10.4 | 9.3x10^-6^ |
| Reading | -51.3 | 9.4 | 5.8x10^-8^ |
| Bury | -29.3 | 9.5 | 2.1x10^-3^ |
| Newcastle | -9.7 | 9.3 | 0.29 |
| Leeds | -35.9 | 9.0 | 7.3x10^-5^ |
| Bristol | -48.3 | 9.1 | 1.0x10^-7^ |
| Barts | 16.3 | 13.6 | 0.23 |
| Nottingham | -49.5 | 9.4 | 1.2x10^-7^ |
| Sheffield | -40.3 | 9.5 | 2.4x10^-5^ |
| Liverpool | -17.5 | 9.4 | 0.06 |
| Middlesborough | -26.1 | 10.1 | 9.7x10^-3^ |
| Hounslow | -23.6 | 10.1 | 0.02 |
| Croydon | -35.7 | 10.0 | 3.4x10^-4^ |
| Birmingham | -30.2 | 10.0 | 2.5x10^-3^ |
| Swansea | -32.0 | 19.6 | 0.10 |
| Wrexham | 1.9 | 34.9 | 0.96 |

^a^SE = standard error

^b^Categorical variables for which the data is presented as change in birthweight in grams compared to the referent category

^c^eGFR = estimated glomerular filtration rate

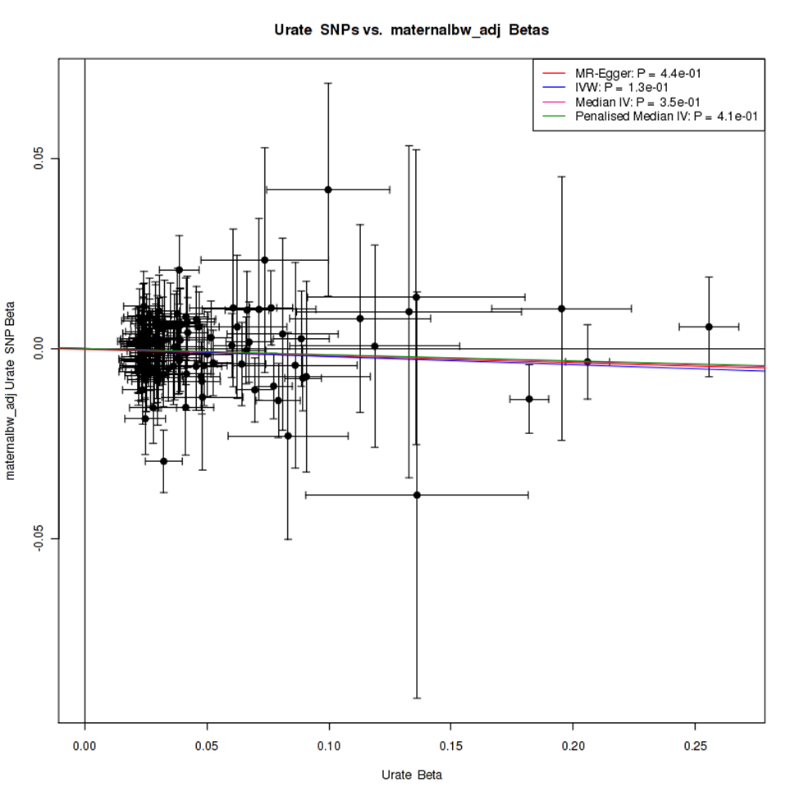

**Supplementary Figure S1. Univariate Mendelian randomization (MR) plot of the estimated causal effect of urate on offspring birth weight using 114 independent urate-associated SNPs.** The x-axis represents the SNP-exposure association while the y-axis represents the SNP-outcome association. Inverse variance weighted (IVW), MR-Egger, weighted median (Median IV), and penalised weighted median (Penalised Median IV) estimates for pleiotropy investigation and estimate comparison.
