## Supplementary Note for "Investigating a possible causal relationship between maternal serum urate concentrations and offspring birthweight: A Mendelian randomization study"

**Estimating Change in Birthweight if Urate was Completely Mediated by Systolic Blood Pressure**

1 umol/L urate = 0.03mmHg systolic blood pressure (SBP)

10mmHg SBP = -0.148 SD birthweight (BW), therefore, 0.0004 SD BW per 1 umol/L

0.0004 multiplied by 1 SD of BW at either 484g or 477g (in Exeter Family Study of Childhood Health) = 0.19g per 1 umol/L urate

To get the per SD of urate effect, 0.19g multiplied by 1 SD of urate at 65 umol/L = 12.35 g BW per 1 SD urate

When taking into account the effect of SBP on BW, there is a 12.35g change in BW per 1 SD change in urate.
